# Joint Association of Polygenic Risk and Social Determinants of Health with Coronary Heart Disease in the United States

**DOI:** 10.1101/2024.01.10.24301105

**Authors:** Kristjan Norland, Daniel J. Schaid, Mohammadreza Naderian, Jie Na, Iftikhar J. Kullo

## Abstract

**Background:** The joint effects of polygenic risk and social determinants of health (SDOH) on coronary heart disease (CHD) in the United States are unknown.

**Methods:** In 67,256 All of Us (AoU) participants with available SDOH data, we ascertained self-reported race/ethnicity and calculated a polygenic risk score for CHD (PRS_CHD_). We used 90 SDOH survey questions to develop an SDOH score for CHD (SDOH_CHD_). We assessed the distribution of SDOH_CHD_ across self-reported races and US states. We tested the joint association of SDOH_CHD_ and PRS_CHD_ with CHD in regression models that included clinical risk factors.

**Results:** SDOH_CHD_ was highest in self-reported black and Hispanic people. Self-reporting as black was associated with higher odds of CHD but not after adjustment for SDOH_CHD_. Median SDOH_CHD_ values varied by US state and were associated with heart disease mortality. A 1-SD increase in SDOH_CHD_ was associated with CHD (OR=1.36; 95% CI, 1.29 to 1.46) and incident CHD (HR=1.73; 95% CI, 1.27 to 2.35) in models that included PRS_CHD_ and clinical risk factors. Among people in the top 20% of PRS_CHD_, CHD prevalence was 4.8% and 7.8% in the bottom and top 20% of SDOH_CHD_, respectively.

**Conclusions:** Increased odds of CHD in self-reported black people are likely due to higher SDOH burden. SDOH and PRS were independently associated with CHD in the US. Our findings emphasize the need to consider both PRS and SDOH for equitable disease risk assessment.

## Introduction

Both genetic and environmental factors contribute to susceptibility to common diseases. Additive genetic liability can be estimated with polygenic risk scores (PRS) (1) but our knowledge of how environmental variables add to or modify polygenic risk is rudimentary (2). Social determinants of health (SDOH) are the environmental conditions that influence health outcomes (3). In the United States (US), SDOH contribute to many health disparities, including mortality from cardiovascular disease (4), and are associated with self-reported race (5).

Coronary heart disease (CHD) is the leading cause of mortality in the US (6,7) and has well-known genetic and environmental susceptibility factors (8). It is also preventable; risk factors are identifiable and 10-year risk can be estimated from clinical risk prediction equations (9). These equations include race as an input variable but not PRS or SDOH. The inclusion of race in clinical algorithms is increasingly under scrutiny as it may contribute to systematic racism in medicine (10). Race is a social, not biological, construct, and SDOH are hypothesized to confound the association of self-reported race with disease risk (11,12). Socioeconomically disadvantaged groups are more often exposed to CHD risk factors such as smoking, have increased stress and reduced access to formal education and medical care, and often live and work in poor conditions (13). Although many other complex diseases fulfill similar criteria, CHD is ideal to study how PRS and SDOH jointly affect the risk of common complex diseases.

Incorporating SDOH information into electronic health records (EHRs) is a relatively recent development, highlighting the need for large cohorts with self-reported SDOH data. The All of Us (AoU) Research Program aims to build the most diverse health dataset in the US, allowing for the integration of SDOH, genetic, and EHR data (14). SDOH data in AoU cover the five SDOH domains defined by *Healthy People 2030* (HP-30). These include 1) economic stability, 2) education access and quality, 3) healthcare access and quality, 4) neighborhood and built environment, and 5) social and community context (15–17).

Recent advances in PRS have raised the prospect of clinical implementation (18,19). In a previous study, we described the variable performance of PRS_CHD_ in three major racial/ethnic groups in the US (20). However, it is unclear whether there are additive or interactive effects between PRS and SDOH on CHD. In this study, we developed an SDOH score for CHD (SDOH_CHD_) in AoU and compared its distribution in different groups of self-reported race and across US states. We tested whether SDOH confound the association of self-reported race with CHD. Finally, we jointly modeled SDOH_CHD_ and PRS_CHD_ in different models of CHD that included clinical risk factors.

## Methods

### Study cohort

Of 245,394 whole-genome sequenced people in the AoU Research Program v7, we analyzed 67,256 who had responded to the AoU SDOH survey. We ascertained demographic factors, CHD, conventional risk factors for CHD (hypertension, type 2 diabetes [T2D], body mass index [BMI], smoking status), medication use (statins and antihypertensives), and other atherosclerotic cardiovascular diseases (ASCVD) from EHR data. We defined CHD by diagnosis and procedure codes (complete definitions for CHD and its risk factors are provided in the Supplementary Appendix). We defined CHD as any code in the EHR and incident CHD as the presence of any code ≥1 month after responding to the SDOH study. We only considered incident CHD events in participants free of CHD at the time of the SDOH study.

### Social determinants of health

We used 81 SDOH questions from the AoU SDOH survey related to loneliness, perceived discrimination, food insecurity, housing instability and quality, perceived stress, daily spiritual experiences, and religious service attendance (Table S1) (16). We included 9 SDOH questions from previous AoU surveys that were not included in the SDOH survey (to avoid duplication of questions) (16). These were related to education and income level, employment and marital status, home ownership and housing instability, and health literacy (Table S1). 81 of the questions were Likert-type or continuous and showed patterns of moderate clustering (Figure S1). We created binary variables out of other questions. In total, we derived 124 variables from the 90 SDOH questions. Further details on the SDOH variables are provided in the Supplementary Appendix.

### Whole-genome sequence data and polygenic risk score for CHD

We used the short-read whole-genome sequencing (srWGS) data, specifically the ACAF-thresholded and filtered PLINK files (variants with population-specific allele frequency <1% or population-specific allele count <100). We used genetic principal components (PCs) and genetic ancestry group predictions provided by the AoU team (see Supplementary Appendix). We used a recent polygenic risk score for CHD (PRS_CHD_) developed from multi-ancestry GWAS summary statistics for CHD (21). PRS_CHD_ was developed using PRS-CSx (22) with 1.3 million variants. When matching variants, we removed ambiguous SNPs and computed the PRS using *plink2*.

### Population descriptors

We assigned participants to major genetic ancestry groups (AFR, AMR, EAS, EUR, MID, and SAS) (23). We used self-reported race and ethnicity from The Basics survey. Due to small group sizes, we binned those who self-reported as multiracial, Middle Eastern and North African (MENA), Native Hawaiian and Pacific Islander (NHPI), or none indicated/prefer not to answer/skip/none of these into a separate category (“other”).

### Statistical analyses

We used the All of Us Researcher Workbench, R version 4.2.2.

We split the study cohort into training and test sets (50/50 split). To impute missing survey responses, we used the R package *missRanger*, which combines random forest imputation with predictive mean matching. To avoid leaking test data into the training set, we first imputed the training set and then the test set separately after combining it with the imputed training set (Supplementary Appendix).

To develop the SDOH_CHD_, we fitted a logistic lasso regression model in the training set with CHD as outcome and potential predictor variables age, age^2^, sex, an indicator variable for self-reported ethnicity, 10 genetic PCs, and 124 SDOH variables. We standardized all variables before fitting the model. We tuned the penalty parameter over a grid of 50 levels, using 10-fold cross-validation repeated ten times, minimizing the log loss. To compute SDOH_CHD_, we standardized SDOH variables according to the means and standard deviations in the training set, multiplied them with corresponding model coefficients, and summed them together.

To test whether the association of self-reported race and CHD was confounded by SDOH_CHD_, we fitted four different logistic regression models that included indicator variables for self-reported races and different sets of covariates: i) age and sex, ii) clinical risk factors, iii) clinical risk factors and PRS_CHD_, and iv) clinical risk factors, PRS_CHD_ and SDOH_CHD_. To test for heterogeneity in the associations of SDOH_CHD_/PRS_CHD_ with CHD between genetic ancestry groups, we fitted logistic regression models of CHD that included SDOH_CHD_/PRS_CHD_, indicator variables for genetic ancestry groups, interaction terms for the two, as well as age, sex, and 10 PCs.

We tested for association between median SDOH_CHD_ by US state of residence and heart disease mortality using a spatial simultaneous autoregressive lag model. Since AoU does not have mortality data, we used mortality rates provided by the Centers for Disease Control and Prevention (CDC) (Supplementary Appendix).

To evaluate SDOH_CHD_ and PRS_CHD_ in the test set, we fitted logistic regression models of CHD that included the scores separately and jointly with interaction. We included two types of models; one included age, sex, and 10 genetic PCs as covariates (basic model), and the other additionally included clinical risk factors (clinical risk factor model; hypertension, T2D, BMI, smoking status, and statin and antihypertensive medications). We standardized PRS_CHD_ within genetic ancestry groups. We used Cox regression to test the association between SDOH_CHD_, PRS_CHD_, and incident CHD. We used age as the time scale and censored participants at their events or on 2022/07/01 (the date of the last observed EHR record), whichever was first. We excluded participants with CHD <1 month after responding to the SDOH survey. We assessed the proportional hazard assumptions for our models (Table S2).

## Results

Of 245,394 participants with available whole-genome sequence data in version 7 of the All of Us (AoU) Research Program, 67,256 responded to the SDOH survey between 2021/11/01 and 2022/06/30. 43,791 (65.1%) were female, the mean age was 59 years (SD, 16), and 2,735 (4.1%) had coronary heart disease (CHD, Table 1). 77.8% self-reported as white, 7.9% as black, and 9% as Hispanic.

**Table 1.** Demographic characteristics of AoU cohort (n=67,256), stratified by CHD status at the time of the SDOH survey.

| **Characteristics at the time of the survey** | **No CHD**<br>**(n=64,521)** | **CHD**<br>**(n=2,735)** |
| --- | --- | --- |
| Age (years), mean (SD) | 59 (16) | 70 (11) |
| Body mass index (kg/m<sup>2</sup>), mean (SD) | 29.1 (7.0) | 30.5 (6.6) |
|  | **n (%)** | **n (%)** |
| Female at birth | 42,693 (66.2%) | 1,098 (40.1%) |
| **Risk factors and medications** |  |  |
| Hypertension | 18,327 (28.4%) | 2,307 (84.4%) |
| Type 2 diabetes | 6,717 (10.4%) | 1,033 (37.8%) |
| Ever smoker | 22,998 (35.6%) | 1,390 (50.8%) |
| Statins | 10,006 (15.5%) | 1,306 (47.8%) |
| Antihypertensives | 10,470 (16.2%) | 1,188 (43.4%) |
| **Self-reported race** |  |  |
| Asian | 1,640 (2.5%) | 24 (0.9%) |
| Black | 5,071 (7.9%) | 254 (9.3%) |
| White | 50,166 (77.8%) | 2,190 (80.1%) |
| Other | 7,652 (11.9%) | 267 (9.8%) |
| **Self-reported ethnicity** |  |  |
| Hispanic | 5,885 (9.1%) | 159 (5.8%) |
| Not Hispanic | 57,078 (88.5%) | 2,479 (90.6%) |
| Other | 1,588 (2.5%) | 97 (3.5%) |
| **Genetic ancestry group** |  |  |
| AFR | 5,584 (8.7%) | 273 (10.0%) |
| AMR | 5,567 (8.6%) | 140 (5.1%) |
| EUR | 51,244 (79.4%) | 2,285 (83.5%) |
| Other* | 2,126 (3.3%) | 37 (1.4%) |

CHD: coronary heart disease, AFR: African genetic ancestry, AMR: Admixed American genetic ancestry, EUR: European genetic ancestry. \*EAS, MID, and SAS.

We randomized the cohort into 50/50 training and test sets (n=33,628) to develop and test an SDOH score for CHD (SDOH_CHD_). SDOH_CHD_ was a weighted sum of 52 SDOH variables (Figure 1A); higher SDOH_CHD_ values indicated higher CHD-associated SDOH burden (Figure 1B). The variables with the highest weights were related to religiousness/spirituality, employment status, highest education level, perceived discrimination, social support, neighborhood quality, health literacy, and food insecurity (Figure 1A). In the test set, a 1-SD increase in SDOH_CHD_ was associated with CHD in basic and clinical risk factors models (OR=1.59; 95% CI, 1.50 to 1.69, and OR=1.35; 95% CI 1.27 to 1.44, respectively). SDOH_CHD_ was also associated with clinical risk factors for CHD and other ASCVD (Figure S2).

**Figure 1.**
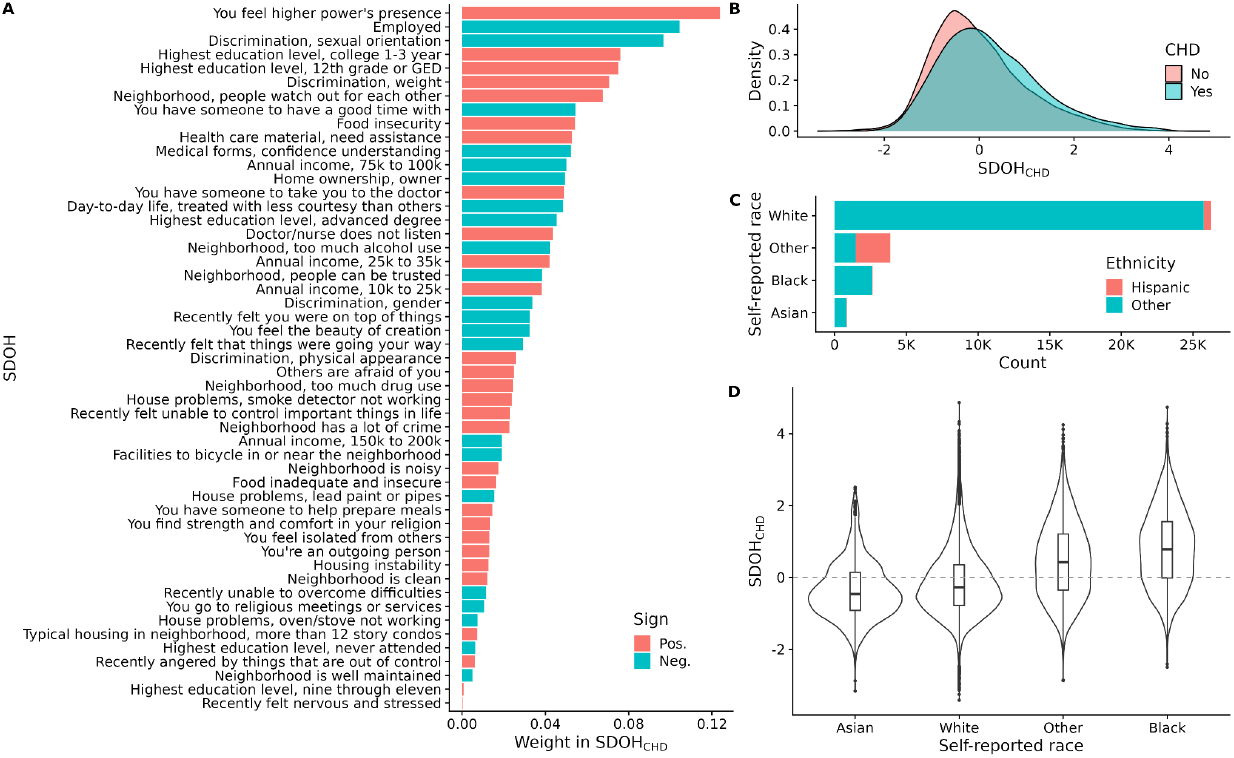
SDOH variables included in SDOH_CHD_, distribution of SDOH_CHD_ stratified by CHD and self-reported race, and the number of participants in the test set, stratified by self-reported race and ethnicity. A) Weights of the 52 SDOH variables included in SDOH_CHD_. The color indicates the sign of the coefficient for the variable. We used simplified labels representing the SDOH variables on the y-axis. B) Distribution of (standardized) SDOH_CHD_ in the test set, stratified by CHD status. C) Number of participants in the test set, stratified by self-reported race and ethnicity. D) Boxplots and violin plots of (standardized) SDOH_CHD_ in the test set, stratified by self-reported race.

In the test set, 26,241 self-reported as white, 2,645 as black, 854 as Asian, and 3,888 as “other”. 61.9% self-reported as Hispanic in the “other” race category, while 1.6-2.6% in the remaining races (Figure 1C). People who self-reported as black or other/Hispanic had the highest SDOH_CHD_ on average (Figure 1D). Self-reporting as black was associated with higher odds of CHD compared to self-reporting as white when adjusting for age and sex (OR=1.70; 95% CI, 1.39 to 2.06) and lower odds of CHD when self-reporting as Asian (OR=0.54; 95% CI, 0.29 to 0.92) (Figure 2A). After adjustment for CHD risk factors and PRS_CHD_, self-reporting as black remained associated with increased odds of CHD (OR=1.28; 95% CI, 1.04 to 1.57) but not after additional adjustment for SDOH_CHD_ (OR=1.05; 95% CI, 0.85 to 1.29).

**Figure 2.**
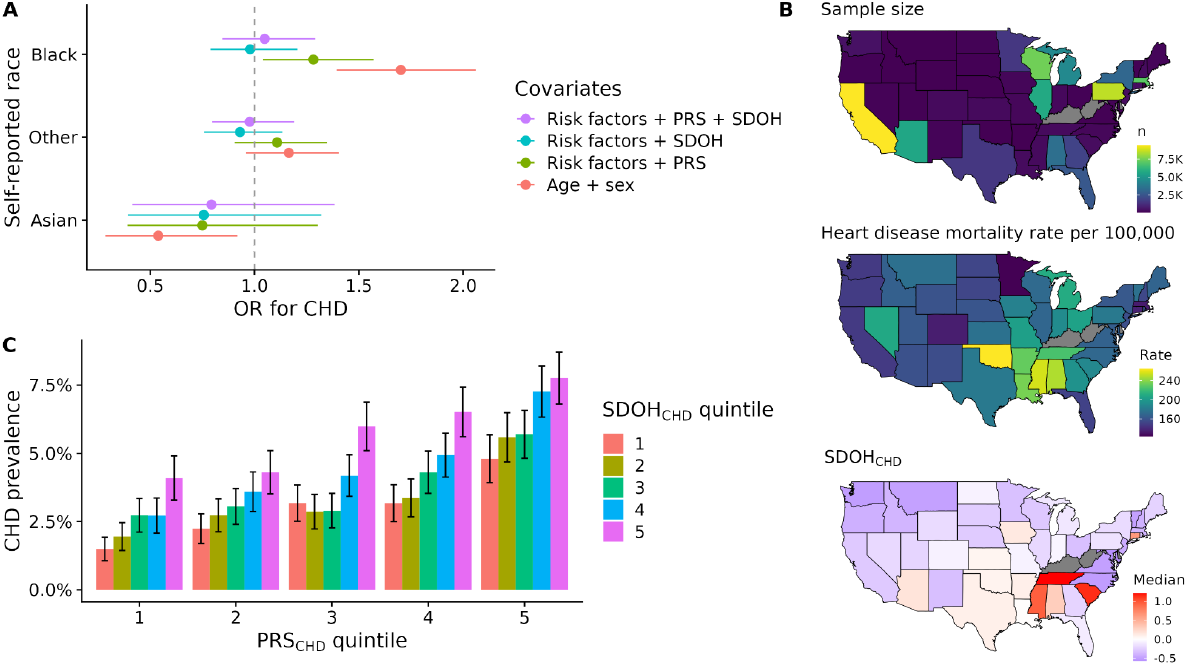
Associations of self-reported race with CHD, geographic variation of SDOH_CHD_ and heart disease mortality by US state, and the distribution of CHD prevalence, stratified by quintiles of SDOH_CHD_ and PRS_CHD_. A) We fitted four logistic regression models of CHD in the test set that included indicator variables for self-reported race (“white” being the reference). We show the OR corresponding to the race indicator variable on the x-axis. Different colors indicate different sets of covariates included in the models. The lines denote 95% CIs. B) Geographic variation of median SDOH_CHD_ values and heart disease mortality by US state. We used the full study cohort (n=67,256) and colored states with <20 participants gray. We standardized SDOH_CHD_ before computing median values. The top choropleth map shows the number of participants by US state, the middle map shows heart disease mortality per 100,000 people by US state, and the bottom map shows median SDOH_CHD_ values by US state. C) Distribution of CHD prevalence in the full study cohort (n=67,256), stratified by quintiles of SDOH_CHD_ and PRS_CHD_. We standardized SDOH_CHD_ in the full study cohort and PRS_CHD_ within genetic ancestry groups. The error bars denote 95% CIs (normal approximations).

We computed median SDOH_CHD_ values by state of residence, which were highest in the Southeast US (SC, TN, MS, AL) and CT (Figure 2B). Median SDOH_CHD_ values were associated with heart disease mortality rates per 100,000 in 2021 as reported by the Centers for Disease Control and Prevention (β=24.8; 95% CI, 7.3 to 42.3) (Figure S3).

We observed high variability in CHD prevalence within quintiles of PRS_CHD_ depending on SDOH_CHD_, and vice versa (Figure 2C, Figure S4). Among people with high genetic risk (fifth quintile of PRS_CHD_), 4.8% (95% CI, 3.9% to 5.7%) and 7.8% (95% CI, 6.8% to 8.7%) had CHD among people with high and low SDOH_CHD_ (fifth and first quintiles of SDOH_CHD_), respectively. CHD prevalence was comparable among people with low genetic risk (first quintile of PRS_CHD_) but high SDOH_CHD_ and those with high genetic risk but low SDOH_CHD_ (4.1% vs 4.8%).

In the test set, the correlation between PRS_CHD_ and SDOH_CHD_ was weak (*r*=0.067; 95% CI, 0.057 to 0.078). There was no significant heterogeneity in the association of SDOH_CHD_ with CHD between genetic ancestry groups (Table S3). For PRS_CHD_, the effect size of PRS_CHD_ was smaller in AFR compared to EUR (*P*<0.001 for the interaction) (Table S4).

Since the effects of SDOH_CHD_ on CHD were not statistically different between genetic ancestry groups, we estimated the joint effects of SDOH_CHD_ and PRS_CHD_ in the full test set. A 1-SD increase in SDOH_CHD_ and PRS_CHD_ was associated with higher odds of CHD when included jointly in a basic model (OR=1.58; 95% CI, 1.49 to 1.68 and OR=1.50; 95% CI, 1.42 to 1.59, respectively) (Table 2, Figure 3A). The effect estimates of SDOH_CHD_ and PRS_CHD_ were similar when included jointly in a model compared to separately (Table 2). Heterogeneity in PRS between ancestry groups likely explained a weak interaction between the scores (OR_int_=0.93; 95% CI, 0.88 to 0.98). In a clinical risk factor model, a 1-SD increase in SDOH_CHD_ and PRS_CHD_ was associated with slightly attenuated odds of CHD compared to the basic model (OR=1.36; 95% CI, 1.28 to 1.46 and OR=1.37; 95% CI, 1.29 to 1.46, respectively).

**Table 2.** Results from logistic and Cox regression models of CHD that included SDOH_CHD_, PRS_CHD_, and different sets of covariates in the test set (n=33,628).

|  |  | CHD |  | Incident CHD |  |
| --- | --- | --- | --- | --- | --- |
|  |  | controls/cases | 32,270/1,358 | participants/events | 32,322/52 |
| Predictor | Covariates | OR per SD (95% CI) | P | HR per SD (95% CI) | P |
| **Models including SDOH<sub>CHD</sub>** |  |  |  |  |  |
| SDOH<sub>CHD</sub> | Basic | 1.59 (1.50 to 1.69) | <0.001 | 2.11 (1.64 to 2.70) | <0.001 |
|  | + Clinical risk factors | 1.35 (1.27 to 1.44) | <0.001 | 1.66 (1.26 to 2.19) | <0.001 |
| **Models including PRS<sub>CHD</sub>** |  |  |  |  |  |
| PRS<sub>CHD</sub> | Basic | 1.53 (1.44 to 1.62) | <0.001 | 1.72 (1.31 to 2.27) | <0.001 |
|  | + Clinical risk factors | 1.37 (1.29 to 1.46) | <0.001 | 1.53 (1.15 to 2.03) | 0.004 |
| **Models including SDOH<sub>CHD</sub> and PRS<sub>CHD</sub> with interaction** |  |  |  |  |  |
| SDOH<sub>CHD</sub> | Basic | 1.58 (1.49 to 1.68) | <0.001 | 2.14 (1.61 to 2.84) | <0.001 |
| PRS<sub>CHD</sub> |  | 1.50 (1.42 to 1.60) | <0.001 | 1.71 (1.27 to 2.34) | <0.001 |
| Interaction term |  | 0.93 (0.88 to 0.98) | 0.01 | 0.90 (0.71 to 1.14) | 0.37 |
| SDOH<sub>CHD</sub> | + Clinical risk factors | 1.36 (1.28 to 1.46) | <0.001 | 1.73 (1.27 to 2.35) | <0.001 |
| PRS<sub>CHD</sub> |  | 1.37 (1.29 to 1.46) | <0.001 | 1.57 (1.15 to 2.14) | 0.005 |
| Interaction term |  | 0.94 (0.89 to 1.00) | 0.05 | 0.89 (0.70 to 1.15) | 0.38 |

The basic covariate model included age, sex, and 10 PCs. The clinical risk factor model included age, sex, 10 PCs, hypertension, T2D, BMI, smoking status, and statin and antihypertensive medications. We standardized SDOH<sub>CHD</sub> within the full test set and PRS<sub>CHD</sub> within each genetic ancestry group.

**Figure 3.**
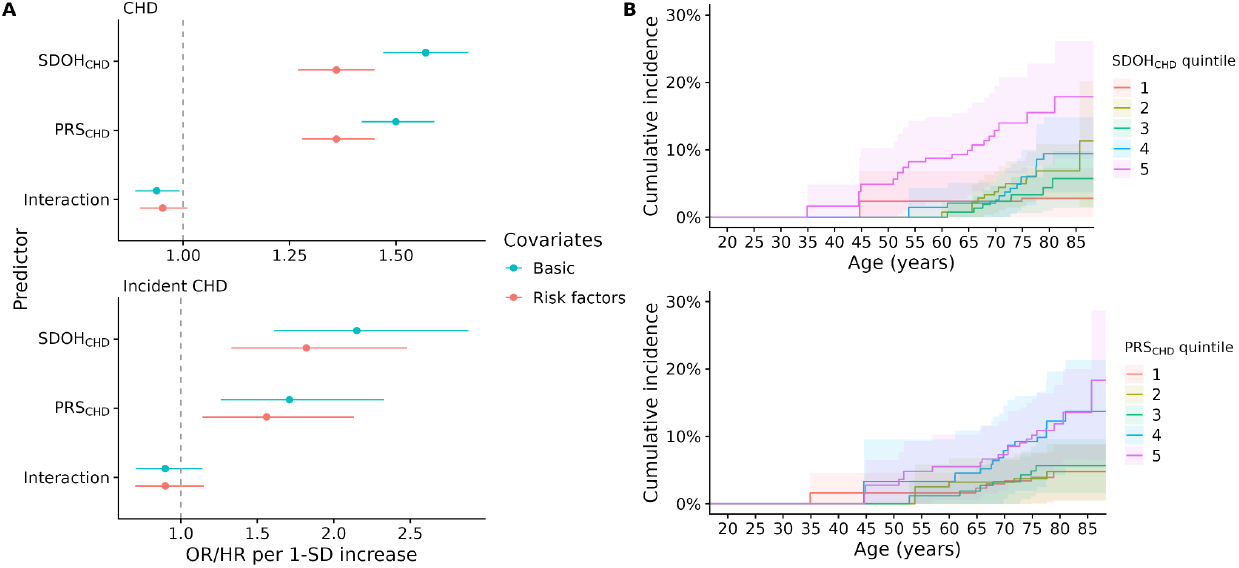
Estimates from regression models of CHD and cumulative incidence of CHD in the test set. A) Results from logistic and Cox regression models of CHD (top) and incident CHD (bottom) that included SDOH_CHD_, PRS_CHD_, an interaction term for the scores, and different sets of covariates (indicated by the colors). B) Cumulative incidence of CHD over a median follow-up of 214 days (IQR, 88) from the time of the SDOH survey, stratified by quintiles of SDOH_CHD_ (top) and quintiles of PRS_CHD_ (bottom).

Over a median follow-up time of 214 days (IQR, 88) from the time of the SDOH survey, we observed 52 incident CHD events in 32,322 participants (excluding 1,306 with CHD diagnosis before the SDOH survey). A 1-SD increase in SDOH_CHD_ and PRS_CHD_ was associated with comparable increased risk of CHD in a basic model (HR=2.14; 95% CI, 1.61 to 2.84 and HR=1.71; 95% CI, 1.27 to 2.34, respectively) and a clinical risk factor model (HR=1.73; 95% CI, 1.27 to 2.35 and HR=1.57; 95% CI, 1.15 to 2.14, respectively) (Table 2, Figure 3).

## Discussion

To address the gap in our knowledge of how environmental factors add to or modify the polygenic risk of CHD, we used data from 67,256 whole-genome sequenced individuals who responded to the SDOH survey in the All of Us (AoU) Research Program. We calculated a multi-ancestry polygenic risk score for CHD (PRS_CHD_) (21) and created an SDOH score for CHD (SDOH_CHD_). SDOH_CHD_ was highest in self-reported black and Hispanic people, and self-reporting as black was associated with increased odds of CHD but not after adjustment for SDOH_CHD_. Median SDOH_CHD_ values by state were associated with heart disease mortality and were highest in the South. Among people in the top 20% of PRS_CHD_, we observed 40% lower CHD prevalence in the first quintile of SDOH_CHD_ compared to the fifth quintile of SDOH_CHD_. A 1-SD increase in SDOH_CHD_ and PRS_CHD_ was associated with comparable odds (risk) of (incident) CHD in models that included clinical risk factors.

Self-reporting as black was associated with higher odds of CHD after adjustment for PRS_CHD_ and clinical risk factors but not after additional adjustment for SDOH_CHD_. This is consistent with results from a prior study showing that racial differences in premature all-cause mortality are confounded by SDOH (11). Recently, new race-free prediction equations for 10- and 30-year risk estimates of cardiovascular disease were proposed, taking into account SDOH to more equitably estimate and address risk (24). Our observational study cannot establish causality but our findings suggest that SDOH confounds the association between race and CHD.

In the test set, a 1-SD increase in both SDOH_CHD_ and PRS_CHD_ was associated with a comparable increase in the odds of CHD when jointly included in models with clinical risk factors. Although we developed SDOH_CHD_ from mostly prevalent CHD, it was also associated with incident CHD events. These results highlight the importance of jointly modeling SDOH and PRS when estimating CHD risk. Our approach extends previous efforts to create SDOH scores for CVD (25,26) and is likely applicable to other common diseases. For example, a recent study showed that T2D prevalence and BMI were influenced by PRS and SDOH in the US and the UK (27). However, since SDOH_CHD_ was specifically developed for CHD, it is different from other commonly used social and economic deprivation indices based on area-level socioeconomic measures.

A strength of our study is a relatively large and diverse cohort representing the US population. The AoU dataset covers all five domains of HP-30 and provides self-reported SDOH measures that cannot be inferred from area-level data. These include SDOH that are known to play a role in CHD, such as employment status and educational attainment, health literacy, and food insecurity (28–31). Our analysis also suggests that other SDOH affect the risk of developing CHD, including perceived discrimination, religiousness/spirituality, perceived stress, neighborhood quality, and social support.

Our study had several limitations. First, the cohort consisted mostly of self-reported white people (78%), which may have influenced the SDOH score. Second, the AoU SDOH survey had patterns in survey nonresponse among people who were non-white and had lower education and income. There was item nonresponse due to racial identity, educational attainment, income level, and age (16). By imputing the missing data, we possibly reduced bias due to item nonresponse. Third, our results need external validations. We are not aware of comparable cohorts with both genetic and extensive SDOH data. Our work highlights the need for systematic collection of SDOH data in other cohorts. Fourth, SDOH_CHD_ is not a typical social deprivation index since the outcome of interest was CHD. Our cross-sectional study precludes causal inferences as some SDOH could result from prevalent disease (“reverse causality”). An ideal SDOH risk score for CHD would be one developed in people without cardiovascular disease at the time of the survey using incident CHD events. Future studies are needed to explore this approach.

Although SDOH_CHD_ was associated with incident CHD, the follow-up time was short, and the number of events was modest. Finally, causal variables that were not accounted for may have confounded associations with SDOH; many variables related to religiousness/spirituality had a positive weight in SDOH_CHD_ but such measures have previously been associated with lower CVD risk, notably among African Americans (32).

Despite the above limitations, our study provides insights into how PRS and SDOH jointly contribute to CHD in the US. The findings could have implications for public health policy and more equitable CHD risk prediction at the individual level. Reducing the SDOH burden with policy changes could decrease heart disease incidence and racial disparities (33). At the individual level, race should be replaced with SDOH in risk prediction equations and joint modeling of PRS and SDOH scores could improve the accuracy of risk estimates.

In conclusion, the burden of SDOH is highest in self-reported black and Hispanic people in the US. Self-reported race was associated with CHD but not after adjustment for SDOH_CHD_. SDOH_CHD_ and PRS_CHD_ were associated with both prevalent and incident CHD after adjustment for clinical risk factors. The effects of PRS and SDOH were additive, implying that joint modeling of the two could improve accuracy and equality in disease risk assessment.

## Data availability

How to request access to data in the All of Us Research Program: https://www.researchallofus.org/.

