## Supplementary material for "Joint Association of Polygenic Risk and Social Determinants of Health with Coronary Heart Disease in the United States"

### Table of contents

**List of investigators** **2**

**Supplementary Methods** **3**

Definitions of coronary heart disease, its risk factors, and other ASCVD 3

Social determinants of health 3

Prediction of genetic ancestry in AoU 4

Imputation of SDOH responses and other variables 4

Geographic variation in $SDOH_{CHD}$ and association with heart disease mortality rates 4

**Supplementary Figures** **6**

Figure S1. Correlations of Likert-type SDOH variables. 6

Figure S2. Associations between $SDOH_{CHD}$ and CHD, its risk factors, and other ASCVD. 7

Figure S3. Median $SDOH_{CHD}$ values by US state and heart disease mortality rates in 2021 provided by the Centers for Disease Control and Prevention (CDC). 8

Figure S4. Distribution of CHD prevalence in the full study cohort, stratified by quintiles of $SDOH_{CHD}$ and $PRS_{CHD}$. 9

**Supplementary Tables** **10**

Table S1. SDOH variables used in the analysis. 10

Table S2. Assessment of the proportional hazards assumption for the two Cox regression models. 14

Table S3. Test for heterogeneity in the effect of $SDOH_{CHD}$ on CHD between genetic ancestry groups. 16

Table S4. Test for heterogeneity in the effect of $PRS_{CHD}$ on CHD between genetic ancestry groups. 17

#### List of investigators

| Investigator | Affiliations |
| --- | --- |
| Kristjan Norland | Department of Cardiovascular Medicine, Mayo Clinic, Rochester, MN, USA |
| Daniel J. Schaid | Department of Quantitative Health Sciences, Mayo Clinic, Rochester, MN, USA |
| Mohammadreza Naderian | Department of Cardiovascular Medicine, Mayo Clinic, Rochester, MN, USA |
| Jie Na | Department of Quantitative Health Sciences, Mayo Clinic, Rochester, MN, USA |
| Iftikhar J. Kullo | Department of Cardiovascular Medicine, Mayo Clinic, Rochester, MN, USA<br>Gonda Vascular Center, Mayo Clinic, Rochester, MN, USA |

#### Supplementary Methods

##### Definitions of coronary heart disease, its risk factors, and other ASCVD

We defined CHD by ≥1 diagnostic codes (ICD-9 codes 410.X, 411.0, 412.X, 429.79 and ICD-10 codes I21.X, I22.X, I23.X, I24.1, I25.2) or ≥1 procedural codes (CPT codes 92920, 92921, 92924, 92925, 92928, 92929, 92933, 92934, 92937, 92938, 92941, 92943, 92944, 92980, 92981, 92982, 92984, 92995, 92996, 92973, 92974, 33510, 33511, 33512, 33513, 33514, 33516, 33517, 33518, 33519, 33521, 33522, 33523, 33533, 33534, 33535, 33536, 62267, 50546; ICD-9 procedure codes 00.66, 36.0X, V45.82, 36.1X, 36.2, V45.81; ICD-10 procedure codes 0270X, 0271X, 0272X, 0273X, 02C0X, 02C1X, 02C2X, 02C3X, 3E07017, 3E070PZ, 3E07317, 3E073PZ, Z95.5, Z98.61, 0210X, 0211X, 0212X, 0213X, Z95.1)

Clinical risk factors: We defined hypertension as the presence of any of the following codes on two separate days: 401, 401.0, 401.9, 402.X, 403.X, 404.X, I10.X, I11.X, I12.X, I13.X, I15.X, I16.X. We defined type 2 diabetes as the presence of any of the following codes on two separate days 250.X, E08.X, E09.X, E10.X, E11.X, E12.X, E13.X. For each participant, we took the mean over all available BMI measurements. We imputed missing BMI values for 996 participants. We defined smoking status (ever smoker) by the "Have you smoked at least 100 cigarettes in your entire life?" question from the AoU Lifestyle survey.

##### Social determinants of health

We analyzed SDOH questions from three AoU surveys: The Basics, Overall Health, and SDOH (Table S1) We analyzed SDOH questions with a completion rate >0.75 (mean completion rate=0.97; excluded one question with high missingness conditional on not speaking English). We converted invalid question responses (e.g. question skipped, don't know, prefer not to answer, other) to NA and imputed those (see Imputation of SDOH responses and other variables). We analyzed Likert-type (e.g. strongly disagree to strongly agree) as continuous. We converted multiple-choice questions (related to perceived discrimination and house problems) to binary variables. We converted other questions to binary variables (e.g. highest education and income levels). We defined employed as employed for wages/student/retired vs. others. We defined marital status as married/living with a partner vs. others.

#### Prediction of genetic ancestry in AoU

The AoU team computed ancestry predictions for all srWGS samples. They used a high-quality set of sites. They trained a random forest classifier on a training set of HGDP and 1000 Genomes sample variants on the autosomal exome, obtained from gnomAD. They generated the first 16 PCs of the training sample genotypes. They used the truth labels from the sample metadata but did not train the classifier on the samples labeled as "Other". To predict the genetic ancestry of "Other", they used probabilities on the other ancestries. To predict the genetic ancestry of AoU samples, they projected AoU samples onto the PC-space of the training data and applied the classifier. More details on the method can be found in the AoU QC Report v7.

#### Imputation of SDOH responses and other variables

We imputed missing SDOH responses and BMI values with the R package *missRanger*, which combines random forest imputation with predictive mean matching. We tried to impute all SDOH variables except the multiple choice questions (perceived discrimination, house problems), which we assumed to be zero if none was indicated. We converted the Likert-type variables to integers before imputing but converted other variables into binary variables after the imputation step. We also included age, sex, 10 PCs, and self-reported ethnicity in the imputation model. We converted invalid survey responses to NA before the imputation (question skipped, don't know, prefer not to answer). We used the parameters "pmm.k = 3, num.trees = 75, sample.fraction = 0.2", where "pmm.k" is the number of candidate non-missing values to sample from in the predictive mean matching steps, "num.trees" is the number of trees, and "sample.fraction" is the size of the bootstrap samples. To avoid leaking test data into the training set, we first imputed the training set and then the test set separately after combining it with the imputed training set. The mean best average OOB imputation error in the training set was 0.108 and 0.093 in the test set.

#### Geographic variation in $SDOH_{CHD}$ and association with heart disease mortality rates

We obtained heart disease mortality data from the Centers for Disease Control and Prevention (CDC) website on 2023/09/15. Heart disease mortality was defined by ICD-10 underlying cause-of-death codes I00–I09, I11, I13, and I20–I51. We computed median $SDOH_{CHD}$ values by state (state of residency). We excluded states with <20 participants (AoU policy). We used a spatial simultaneous autoregressive lag model implemented in the R package *spatialreg* to account for the spatial autocorrelation in the mortality data. We downloaded a US state boundary file from https://www.census.gov/geographies/mapping-files/time-series/geo/carto-boundary-file.html (cb_2018_us_state_500k.zip). We used poly2nb (from the R package *spdep*) to construct a neighbours list from the boundary file.

#### Supplementary Figures

##### Figure S1. Correlations of Likert-type SDOH variables.

Spearman’s correlations of Likert-type SDOH questions. We used Spearman’s rank correlation as a distance metric and clustered the questions hierarchically with complete linkage. We used simplified labels for the sources/constructs that the questions belong to.

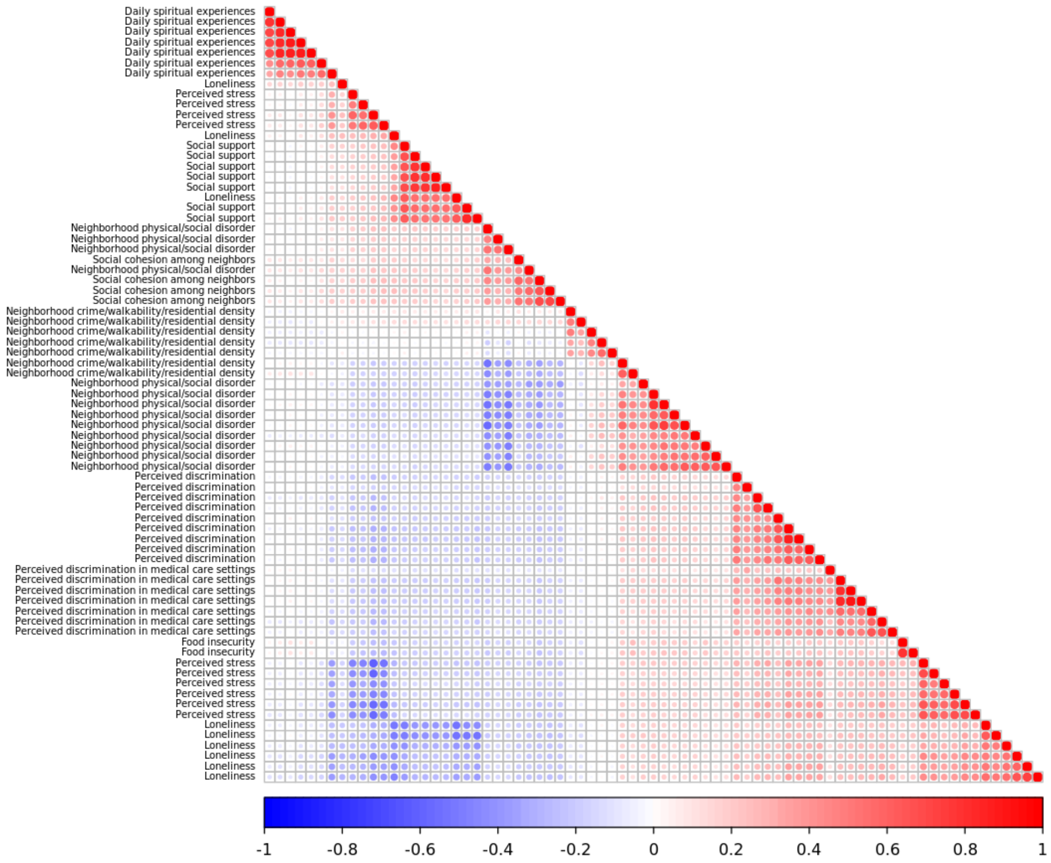

**Figure S2. Associations between $SDOH_{CHD}$ and CHD, its risk factors, and other ASCVD.**

Odds ratios per 1-SD increase in $SDOH_{CHD}$ for CHD, its risk factors, and other ASCVD. We included the traits separately in logistic regression models and adjusted for age, sex, and 10 PCs.

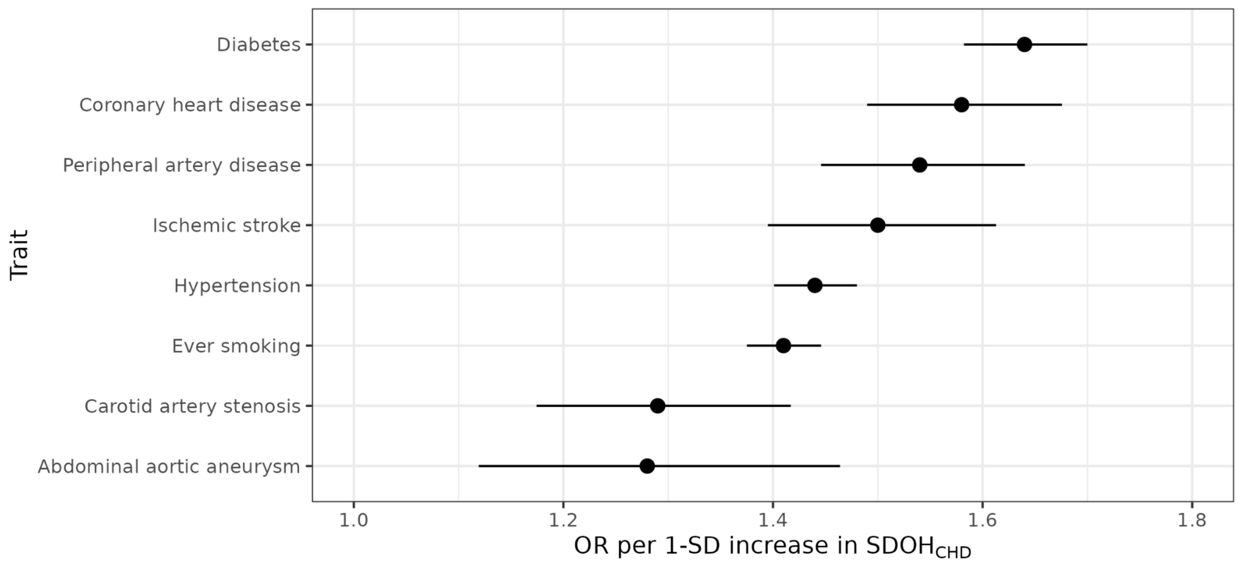

**Figure S3. Median $SDOH_{CHD}$ values by US state and heart disease mortality rates in 2021 provided by the Centers for Disease Control and Prevention (CDC).**

The relationship between median $SDOH_{CHD}$ values by US state with heart disease mortality. The blue line represents a fit from a generalized additive model (GAM). The lines represent 95% CIs.

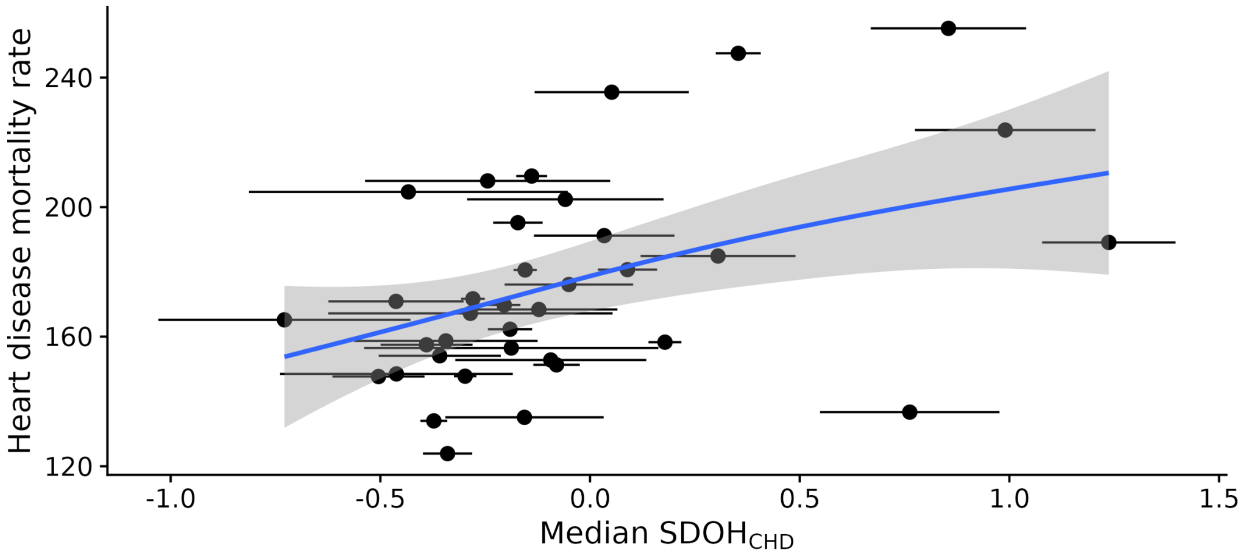

**Figure S4. Distribution of CHD prevalence in the full study cohort, stratified by quintiles of $SDOH_{CHD}$ and $PRS_{CHD}$.**

The error bars indicate 95% CIs.

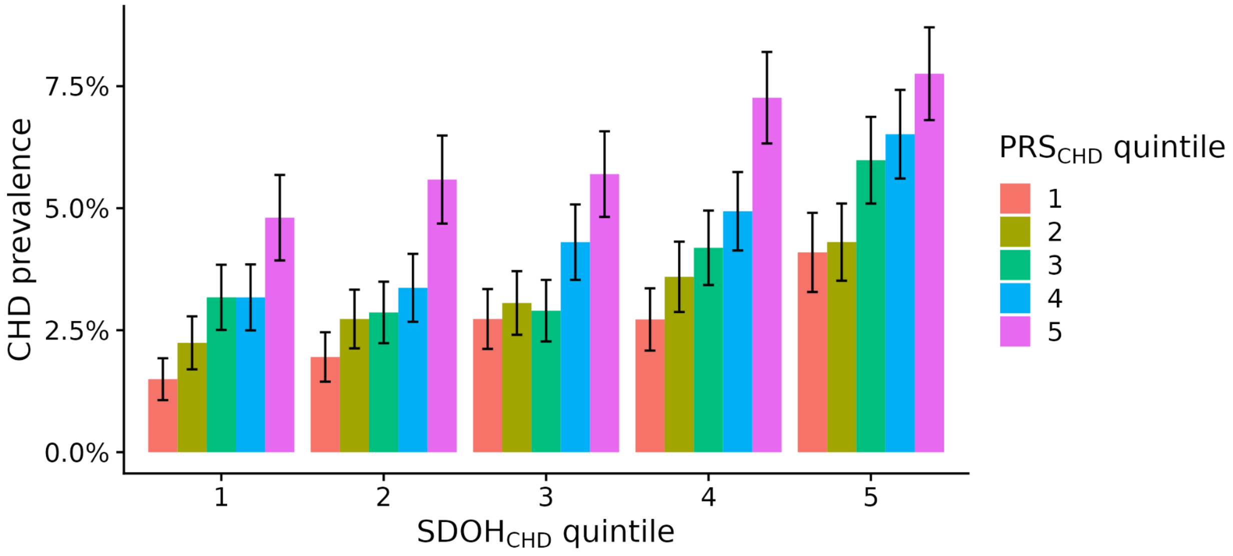

#### Supplementary Tables

##### Table S1. SDOH variables used in the analysis.

The 124 SDOH variables that we included in the development of $SDOH_{CHD}$. We provide a distribution of the responses in the last column. Higher values indicate more agreement for the Likert-type variables (responses starting with "1").

| Question | Survey | Missing rate (before imputation) | Responses (after imputation) |
|---|---|---|---|
| How often do you feel isolated from others? | SDOH | 0.0321 | 1=24306; 2=22158; 3=16442; 4=4350 |
| How often do you feel lack companionship? | SDOH | 0.014 | 1=24726; 2=19231; 3=16474; 4=6825 |
| How often do you feel that you are an outgoing person? | SDOH | 0.0192 | 1=2381; 2=8658; 3=29764; 4=26453 |
| How often do you feel God's (or a higher power's) presence? | SDOH | 0.0141 | 1=12204; 2=8143; 3=8924; 4=6322; 5=7987; 6=12817; 7=10859 |
| How often do you feel that there is no one you can turn to? | SDOH | 0.0204 | 1=34808; 2=19107; 3=9956; 4=3385 |
| How often do you have someone who understands your problems? | SDOH | 0.0728 | 1=8691; 2=13812; 3=23395; 4=21358 |
| How much you agree or disagree that your neighborhood is safe? | SDOH | 0.0256 | 1=1712; 2=3954; 3=35148; 4=26442 |
| How often do you feel that you are unhappy being so withdrawn? | SDOH | 0.0223 | 1=31290; 2=20000; 3=12769; 4=3197 |
| How much you agree or disagree that your neighborhood is clean? | SDOH | 0.0515 | 1=3020; 2=5968; 3=29121; 4=29147 |
| How much you agree or disagree that your neighborhood is noisy? | SDOH | 0.0243 | 1=28398; 2=25561; 3=10128; 4=3169 |
| How often do you have someone to love and make you feel wanted? | SDOH | 0.0845 | 1=7762; 2=9259; 3=16938; 4=33297 |
| How often do you feel that people are around you but not with you? | SDOH | 0.0255 | 1=17958; 2=22355; 3=20877; 4=6066 |
| In the last month, how often have you felt nervous and "stressed"? | SDOH | 0.0344 | 1=8221; 2=17656; 3=24594; 4=10482; 5=6303 |
| In your day-to-day life, how often are you threatened or harassed? | SDOH | 0.0365 | 1=47055; 2=13369; 3=4805; 4=1164; 5=509; 6=354 |
| In your day-to-day life, how often are you called names or insulted? | SDOH | 0.0286 | 1=45144; 2=13347; 3=5812; 4=1552; 5=799; 6=602 |
| How often do you have someone to help you if you were confined to bed? | SDOH | 0.0781 | 1=8164; 2=12487; 3=20191; 4=26414 |
| How often do you fell that you can find companionship when you want it? | SDOH | 0.0195 | 1=2060; 2=4490; 3=19191; 4=41515 |
| How often do you have someone to take you to the doctor if you need it? | SDOH | 0.0622 | 1=6275; 2=10026; 3=19956; 4=30999 |
| In the last month, how often have you felt that you were on top of things? | SDOH | 0.0425 | 1=1640; 2=5669; 3=17288; 4=24402; 5=18257 |
| How much you agree or disagree that vandalism is common in your neighborhood? | SDOH | 0.0283 | 1=38619; 2=22483; 3=4453; 4=1701 |
| How often do you have someone to help you with daily chores if you were sick? | SDOH | 0.122 | 1=11888; 2=13053; 3=17411; 4=24904 |
| In your day-to-day life, how often do people act as if they are afraid of you? | SDOH | 0.0554 | 1=47219; 2=10792; 3=5905; 4=1905; 5=818; 6=617 |
| How much you agree or disagree that people in your neighborhood can be trusted? | SDOH | 0.025 | 1=910; 2=2934; 3=16940; 4=30547; 5=15925 |
| How much you agree or disagree that there is a lot of crime in your neighborhood? | SDOH | 0.0306 | 1=36733; 2=23972; 3=5080; 4=1471 |
| In your day-to-day life, how often do people act as if they're better than you are? | SDOH | 0.0241 | 1=24715; 2=16820; 3=17361; 4=4401; 5=2074; 6=1885 |
| How much you agree or disagree that there is a lot of graffiti in your neighborhood? | SDOH | 0.0192 | 1=44392; 2=16996; 3=3174; 4=2694 |
| How much you agree or disagree that there is too much drug use in your neighborhood? | SDOH | 0.0462 | 1=36726; 2=22625; 3=6023; 4=1882 |
| In the last month, how often have you been able to control irritations in your life? | SDOH | 0.0273 | 1=2355; 2=4693; 3=16352; 4=23709; 5=20147 |
| In your day-to-day life, how often do people act as if they think you are dishonest? | SDOH | 0.0189 | 1=47728; 2=12762; 3=4397; 4=1237; 5=645; 6=487 |

| | | | |
|---|---|---|---|
| In your day-to-day life, how often do people act as if they think you are not smart? | SDOH | 0.0381 | 1=32885; 2=15956; 3=11741; 4=3563; 5=1752; 6=1359 |
| How much you agree or disagree that people in your neighborhood share the same values? | SDOH | 0.0349 | 1=1330; 2=6725; 3=28351; 4=23125; 5=7725 |
| How much you agree or disagree that you are always having trouble with your neighbors? | SDOH | 0.0207 | 1=43970; 2=21054; 3=1491; 4=741 |
| In your day-to-day life, how often are you treated with less respect than other people? | SDOH | 0.0192 | 1=27442; 2=18183; 3=14354; 4=4143; 5=2038; 6=1096 |
| In your day-to-day life, how often are you treated with less courtesy than other people? | SDOH | 0.0119 | 1=27580; 2=17779; 3=14503; 4=4159; 5=2088; 6=1147 |
| There are sidewalks on most of the streets in my neighborhood. Would you say that you... | SDOH | 0.0203 | 1=14326; 2=4664; 3=2203; 4=8891; 5=37172 |
| How much you agree or disagree that in your neighborhood people watch out for each other? | SDOH | 0.024 | 1=2065; 2=8164; 3=39725; 4=17302 |
| How often do you have someone to prepare your meals if you were unable to do it yourself? | SDOH | 0.103 | 1=9685; 2=12834; 3=17892; 4=26845 |
| How often do you feel God's (or a higher power's) love for you, directly or through others? | SDOH | 0.0243 | 1=13655; 2=7980; 3=7307; 4=7308; 5=9484; 6=12398; 7=9124 |
| In the last month, how often have you been upset because of something that happened unexpectedly? | SDOH | 0.012 | 1=10272; 2=25124; 3=23591; 4=5918; 5=2351 |
| How often do you have someone to turn to for suggestions about how to deal with a personal problem? | SDOH | 0.0663 | 1=7465; 2=12360; 3=21600; 4=25831 |
| The crime rate in my neighborhood makes it unsafe to go on walks at night. Would you say that you... | SDOH | 0.0595 | 1=37338; 2=17675; 3=8563; 4=3680 |
| In the last month, how often have you been angered because of things that were outside of your control? | SDOH | 0.0574 | 1=11182; 2=23900; 3=23212; 4=6623; 5=2339 |
| In the last month, how often have you felt confident about your ability to handle your personal problems? | SDOH | 0.055 | 1=2723; 2=4572; 3=13454; 4=20224; 5=26283 |
| The crime rate in my neighborhood makes it unsafe to go on walks during the day. Would you say that you... | SDOH | 0.116 | 1=53378; 2=9699; 3=2931; 4=1248 |
| In the last month, how often have you felt that you were unable to control the important things in your life? | SDOH | 0.0175 | 1=16561; 2=22562; 3=18891; 4=6193; 5=3049 |
| In the last month, how often have you felt difficulties were piling up so high that you could not overcome them? | SDOH | 0.0933 | 1=24804; 2=21448; 3=13946; 4=4675; 5=2383 |
| How often do you receive poorer service than others when you go to a doctor's office or other health care provider? | SDOH | 0.0287 | 1=40392; 2=19462; 3=6056; 4=606; 5=740 |
| How often are you treated with less respect than other people when you go to a doctor's office or other health care provider? | SDOH | 0.0235 | 1=39270; 2=19809; 3=6757; 4=669; 5=751 |
| It is within a 10-15 minute walk to a transit stop (such as bus, train, trolley, or tram) from my home. Would you say that you... | SDOH | 0.061 | 1=21760; 2=4917; 3=9851; 4=30728 |
| Within the past 12 months, were you worried whether the food you had bought just didn't last and you didn't have money to get more? | SDOH | 0.0122 | 1=61269; 2=4717; 3=1270 |
| How often does a doctor or nurse act as if he or she is afraid of you when you go to a doctor's office or other health care provider? | SDOH | 0.0638 | 1=60587; 2=4919; 3=956; 4=135; 5=659 |
| How often does a doctor or nurse act as if he or she is better than you when you go to a doctor's office or other health care provider? | SDOH | 0.019 | 1=39257; 2=17629; 3=8484; 4=1327; 5=559 |
| Many shops, stores, markets or other places to buy things I need are within easy walking distance of my home. Would you say that you... | SDOH | 0.017 | 1=21039; 2=11766; 3=16878; 4=17573 |
| How often does a doctor or nurse act as if he or she thinks you are not smart when you go to a doctor's office or other health care provider? | SDOH | 0.035 | 1=43642; 2=15047; 3=6702; 4=1025; 5=840 |
| How often do you feel like a doctor or nurse is not listening to what you were saying. when you go to a doctor's office or other health care provider? | SDOH | 0.0223 | 1=26938; 2=21553; 3=15407; 4=2486; 5=872 |
| There are facilities to bicycle in or near my neighborhood, such as special lanes, separate paths or trails, or shared use paths for cycles and pedestrians. Would you say that you... | SDOH | 0.0418 | 1=14081; 2=8079; 3=2144; 4=16310; 5=26642 |
| My neighborhood has several free or low-cost recreation facilities, such as parks, walking trails, bike paths, recreation centers, playgrounds, public swimming pools, etc. Would you say that you... | SDOH | 0.278 | 1=9190; 2=7363; 3=21175; 4=29528 |
| How often do you go to religious meetings or services? | SDOH | 0.136 | 1=17337; 2=22579; 3=7400; 4=4565; 5=9807; 6=5568 |
| How often do you have someone to have a good time with? | SDOH | 0.0852 | 1=7133; 2=13588; 3=21117; 4=25418 |
| How often do you find strength and comfort in your religion? | SDOH | 0.0168 | 1=20166; 2=4275; 3=7174; 4=5801; 5=7809; 6=11833; 7=10198 |
| How often do you desire to be closer to or in union with God (or a higher power)? | SDOH | 0.0179 | 1=13203; 2=8669; 3=8553; 4=7326; 5=7469; 6=13258; 7=8778 |
| How much you agree or disagree that there is too much alcohol use in your neighborhood? | SDOH | 0.0803 | 1=30572; 2=28073; 3=6773; 4=1838 |
| How much you agree or disagree that people around here are willing to help their neighbor? | SDOH | 0.0101 | 1=852; 2=2923; 3=13151; 4=31151; 5=19179 |
| How much you agree or disagree that there are lot of abandoned buildings in your neighborhood? | SDOH | 0.0361 | 1=47361; 2=15666; 3=2616; 4=1613 |
| How much you agree or disagree that people in your neighborhood generally get along with each other? | SDOH | 0.0177 | 1=488; 2=1743; 3=11181; 4=36546; 5=17298 |
| How much you agree or disagree that there are too many people hanging around on the streets near your home? | SDOH | 0.0245 | 1=41122; 2=20477; 3=3887; 4=1770 |
| Within the past 12 months, were you worried whether your food would run out before you got money to buy more? | SDOH | 0.00926 | 1=59455; 2=6116; 3=1685 |
| How much you agree or disagree that people in your neighborhood take good care of their houses and apartments? | SDOH | 0.0216 | 1=3070; 2=3980; 3=34109; 4=26097 |

| | | | |
|---|---|---|---|
| How often do you feel deep inner peace or harmony? | SDOH | 0.0159 | 1=4271; 2=10450; 3=13477; 4=19551; 5=11009; 6=8498 |
| In the last month, how often have you felt that things were going your way? | SDOH | 0.0831 | 1=1934; 2=5245; 3=19316; 4=24774; 5=15987 |
| How often do you feel that you are spiritually touched by the beauty of creation? | SDOH | 0.0184 | 1=5911; 2=7138; 3=9335; 4=12802; 5=17274; 6=14796 |
| In the last month, how often have you found that you could not cope with all the things that you had to do? | SDOH | 0.0219 | 1=19077; 2=22426; 3=17694; 4=5497; 5=2562 |
| In your day-to-day life, how often do you receive poorer service than other people at restaurants or stores? | SDOH | 0.0238 | 1=36031; 2=18754; 3=9196; 4=1948; 5=825; 6=502 |
| How often are you treated with less courtesy than other people when you go to a doctor's office or other health care provider? | SDOH | 0.0165 | 1=39283; 2=19866; 3=6552; 4=687; 5=868 |
| In the last 12 months, how many times have you or your family moved from one home to another? Number of moves in past 12 months: | SDOH | 0.0333 | 0=58898; 1=6648; 2=1008; 3=350; 4=121; 5=55; 6=34; 7=28; >7=114 |
| How often do you feel left out? | SDOH | 0.0245 | 1=18848; 2=25753; 3=18826; 4=3829 |
| **Think about the place you live. Do you have problems with any of the following (check all that apply)?** | SDOH | 0 | |
| Inadequate heat | | | 0=64963; 1=2293 |
| Water leaks | | | 0=62093; 1=5163 |
| No or not working smoke detector | | | 0=65411; 1=1845 |
| Over or stove not working | | | 0=66024; 1=1232 |
| Bug infestation | | | 0=62980; 1=4276 |
| Lead paint or pipes | | | 0=65571; 1=1685 |
| Mold | | | 0=62096; 1=5160 |
| **Discrimination. What do you think is the main reason for these experiences? Select all that apply.** | SDOH | 0 | |
| Your race | | | 0=59281; 1=7975 |
| Your education or income level | | | 0=61197; 1=6059 |
| Your gender | | | 0=52367; 1=14889 |
| Some other aspect of your physical appearance | | | 0=62373; 1=4883 |
| Your age | | | 0=52189; 1=15067 |
| Your height | | | 0=64345; 1=2911 |
| Your sexual orientation | | | 0=65192; 1=2064 |
| Your religion | | | 0=65083; 1=2173 |
| Your ancestry or national origins | | | 0=62161; 1=5095 |
| Your weight | | | 0=61012; 1=6244 |
| In the past 6 months, have you been worried or concerned about NOT having a place to live? | The Basics | 0.00614 | 0=61997; 1=5259 |
| Are you married/living with a partner? | The Basics | 0.0103 | 0=25207; 1=42049 |
| Are you employed for wages/a student/retired? | The Basics | 0.0115 | 0=13063; 1=54193 |
| How confident are you filling out medical forms by yourself? | Overall Health | 0.00559 | 1=344; 2=528; 3=2621; 4=12170; 5=51593 |
| How often do you have someone help you read health-related materials? | Overall Health | 0.0065 | 1=49920; 2=10990; 3=4005; 4=1395; 5=946 |
| How often do you have problems learning about your medical condition because of difficulty understanding written information? | Overall Health | 0.00888 | 1=53519; 2=9391; 3=3243; 4=675; 5=428 |
| **What is the main type of housing in your neighborhood?** | SDOH | 0.0204 | |
| Apartments or condos of 4-12 stories | | 0.0204 | 0=64156; 1=3100 |
| Apartments or condos of more than 12 stories | | 0.0204 | 0=65760; 1=1496 |
| Detached single-family housing | | 0.0204 | 0=23881; 1=43375 |
| Mix of single-family residences and townhouses | | 0.0204 | 0=54767; 1=12489 |
| Townhouses | | 0.0204 | 0=60460; 1=6796 |
| **What is the highest grade or year of school you completed?** | The Basics | 0.00975 | |
| Advanced Degree | | 0.00975 | 0=44096; 1=23160 |
| College Graduate | | 0.00975 | 0=47101; 1=20155 |
| College One to Three | | 0.00975 | 0=51158; 1=16098 |
| Five Through Eight | | 0.00975 | 0=66820; 1=436 |
| Never Attended | | 0.00975 | 0=67246; 1=10 |

| | | | |
|---|---|---|---|
| Nine Through Eleven | | 0.00975 | 0=66244; 1=1012 |
| One Through Four | | 0.00975 | 0=67100; 1=156 |
| Twelve Or GED | | 0.00975 | 0=61027; 1=6229 |
| **Do you own or rent the place where you live?** | The Basics | 0.0156 | |
| Other Arrangement | | | 0=63464; 1=3792 |
| Own | | | 0=20556; 1=46700 |
| Rent | | | 0=50492; 1=16764 |
| **What is your annual household income from all sources?** | The Basics | 0.109 | |
| 10k to 25k | | | 0=60410; 1=6846 |
| 25k to 35k | | | 0=62524; 1=4732 |

**Table S2. Assessment of the proportional hazards assumption for the two Cox regression models.**

Output from cox.zph() for the Cox model with basic covariates:

| Variable | chisq | df | p |
|---|---|---|---|
| sdoh_chd | 0.00367 | 1 | 0.95 |
| prs_chd | 1.34236 | 1 | 0.25 |
| sex | 0.67017 | 1 | 0.41 |
| pc1 | 2.19149 | 1 | 0.14 |
| pc2 | 0.01901 | 1 | 0.89 |
| pc3 | 0.00913 | 1 | 0.92 |
| pc4 | 0.75619 | 1 | 0.38 |
| pc5 | 1.09092 | 1 | 0.3 |
| pc6 | 0.14326 | 1 | 0.71 |
| pc7 | 0.11412 | 1 | 0.74 |
| pc8 | 0.01801 | 1 | 0.89 |
| pc9 | 1.54202 | 1 | 0.21 |
| pc10 | 0.00458 | 1 | 0.95 |
| sdoh_chd:prs_chd | 0.70528 | 1 | 0.4 |
| GLOBAL | 10.56974 | 14 | 0.72 |

Output from cox.zph() for the Cox model that included clinical risk factors:

| | chisq | df | p |
|---|---|---|---|
| sdoh_chd | 1.27E-02 | 1 | 0.91 |
| prs_chd | 1.44E+00 | 1 | 0.23 |
| sex | 9.37E-01 | 1 | 0.33 |
| pc1 | 2.56E+00 | 1 | 0.11 |
| pc2 | 7.18E-02 | 1 | 0.79 |
| pc3 | 6.87E-06 | 1 | 1 |
| pc4 | 5.83E-01 | 1 | 0.45 |
| pc5 | 1.30E+00 | 1 | 0.25 |
| pc6 | 1.16E-01 | 1 | 0.73 |
| pc7 | 1.27E-01 | 1 | 0.72 |
| pc8 | 9.05E-03 | 1 | 0.92 |
| pc9 | 1.99E+00 | 1 | 0.16 |
| pc10 | 1.76E-02 | 1 | 0.89 |
| hypertension | 1.40E+00 | 1 | 0.24 |
| diabetes | 1.80E-01 | 1 | 0.67 |
| bmi | 2.08E-01 | 1 | 0.65 |
| statin | 3.81E-02 | 1 | 0.85 |
| anti_hypertensive | 2.22E-03 | 1 | 0.96 |
| smoker | 3.12E-01 | 1 | 0.58 |
| sdoh_chd:prs_chd | 9.49E-01 | 1 | 0.33 |

| | | | |
|---|---|---|---|
| GLOBAL | 1.46E+01 | 20 | 0.8 |

**Table S3. Test for heterogeneity in the effect of $SDOH_{CHD}$ on CHD between genetic ancestry groups.**

We fitted the logistic regression model CHD ~ sdoh_chd + ancestry_pred + sdoh_chd:ancestry_pred + age + sex + 10 PCs in the test set, where sdoh_chd:ancestry_pred is an interaction term for sdoh_chd and genetic ancestry groups. We standardized $SDOH_{CHD}$ within the test set. We show the effect estimates for terms corresponding to the genetic ancestry groups and $SDOH_{CHD}$.

| Term | OR | P | 95% CI |
|---|---|---|---|
| ancestry_pred_afr | 0.92 | 0.87 | 0.33 to 2.47 |
| ancestry_pred_amr | 0.69 | 0.17 | 0.39 to 1.16 |
| ancestry_pred_eas | 0.51 | 0.6 | 0.05 to 6.57 |
| ancestry_pred_mid | 0.31 | 0.25 | 0.02 to 1.47 |
| ancestry_pred_sas | 0.50 | 0.32 | 0.12 to 1.86 |
| sdoh_chd:ancestry_pred_afr | 1.00 | 0.97 | 0.84 to 1.20 |
| sdoh_chd:ancestry_pred_amr | 0.94 | 0.62 | 0.75 to 1.19 |
| sdoh_chd:ancestry_pred_eas | 1.65 | 0.23 | 0.70 to 3.71 |
| sdoh_chd:ancestry_pred_mid | 3.00 | 0.12 | 0.84 to 14.9 |
| sdoh_chd:ancestry_pred_sas | 1.39 | 0.36 | 0.66 to 2.77 |

**Table S4. Test for heterogeneity in the effect of $PRS_{CHD}$ on CHD between genetic ancestry groups.**

We fitted the logistic regression model CHD ~ prs_chd + ancestry_pred + prs_chd:ancestry_pred + age + sex + 10 PCs in the test set, where prs_chd:ancestry_pred is an interaction term for prs_chd and genetic ancestry groups. We standardized $PRS_{CHD}$ within ancestry groups. We show the effect estimates for terms corresponding to the genetic ancestry groups and $PRS_{CHD}$.

| Term | OR | P | 95% CI |
|---|---|---|---|
| ancestry_pred_afr | 0.56 | 3.00E-01 | 0.19 to 1.62 |
| ancestry_pred_amr | 0.40 | 1.10E-02 | 0.20 to 0.80 |
| ancestry_pred_eas | 0.12 | 1.30E-01 | 0.01 to 2.06 |
| ancestry_pred_mid | 0.65 | 5.10E-01 | 0.13 to 1.95 |
| ancestry_pred_sas | 0.29 | 8.70E-02 | 0.07 to 1.15 |
| prs_chd:ancestry_pred_afr | 0.66 | 3.90E-05 | 0.54 to 0.80 |
| prs_chd:ancestry_pred_amr | 1.05 | 8.00E-01 | 0.74 to 1.49 |
| prs_chd:ancestry_pred_eas | 0.60 | 2.60E-01 | 0.24 to 1.43 |
| prs_chd:ancestry_pred_mid | 0.93 | 9.10E-01 | 0.31 to 3.50 |
| prs_chd:ancestry_pred_sas | 0.80 | 5.30E-01 | 0.39 to 1.61 |
